# COVID-19 severity is associated with immunopathology and multi-organ damage

**DOI:** 10.1101/2020.06.19.20134379

**Authors:** Yan-Mei Chen, Yuanting Zheng, Ying Yu, Yunzhi Wang, Qingxia Huang, Feng Qian, Lei Sun, Zhi-Gang Song, Ziyin Chen, Jinwen Feng, Yanpeng An, Jingcheng Yang, Zhenqiang Su, Shanyue Sun, Fahui Dai, Qinsheng Chen, Qinwei Lu, Pengcheng Li, Yun Ling, Zhong Yang, Huiru Tang, Leming Shi, Li Jin, Edward C. Holmes, Chen Ding, Tong-Yu Zhu, Yong-Zhen Zhang

## Abstract

COVID-19 is characterised by dysregulated immune responses, metabolic dysfunction and adverse effects on the function of multiple organs. To understand how host responses contribute to COVID-19 pathophysiology, we used a multi-omics approach to identify molecular markers in peripheral blood and plasma samples that distinguish COVID-19 patients experiencing a range of disease severities. A large number of expressed genes, proteins, metabolites and extracellular RNAs (exRNAs) were identified that exhibited strong associations with various clinical parameters. Multiple sets of tissue-specific proteins and exRNAs varied significantly in both mild and severe patients, indicative of multi-organ damage. The continuous activation of IFN-I signalling and neutrophils, as well as a high level of inflammatory cytokines, were observed in severe disease patients. In contrast, COVID-19 in mild patients was characterised by robust T cell responses. Finally, we show that some of expressed genes, proteins and exRNAs can be used as biomarkers to predict the clinical outcomes of SARS-CoV-2 infection. These data refine our understanding of the pathophysiology and clinical progress of COVID-19 and will help guide future studies in this area.

## Introduction

Coronaviruses (family *Coronaviridae*) are a diverse group of positive-sense single-stranded RNA viruses with enveloped virions (Cui et al., 2019; Masters and Perlman, 2013). Coronaviruses are well known due to the emergence of Severe Acute Respiratory Syndrome (SARS) in 2002–2003 and Middle East Respiratory Syndrome (MERS) in 2012, both of which caused thousands of cases in multiple countries (Bermingham et al., 2012; Cui et al., 2019; Ksiazek et al., 2003). Coronaviruses naturally infect a broad range of vertebrate hosts including mammals and birds (Cui et al., 2019). As coronavirus primarily target epithelial cells, they are generally associated with gastrointestinal and respiratory infections (Cui et al., 2019; Masters and Perlman, 2013). In addition, they cause hepatic and neurological diseases of varying severity (Masters and Perlman, 2013).

The world is currently experiencing a disease pandemic (COVID-19) caused by a newly identified coronavirus called SARS-CoV-2 (Wu et al., 2020a). At the time of writing, there have been more than 6 million cases of SARS-CoV-2 and over 387,000 deaths globally (WHO, 2020). The disease leads to both mild and severe respiratory manifestations, with the latter prominent in the elderly and those with underlying medical conditions such as cardiovascular and chronic respiratory disease, diabetes, and cancer (Guan et al., 2020). In addition to respiratory syndrome, mild gastrointestinal and/or cardiovascular symptoms as well as neurological manifestations have been documented in hospitalized COVID-19 patients (Mao et al., 2020). Combined, these data point to multiple organ failures, and hence that COVID-19 pathogenesis is complex, especially in patients experiencing severe disease.

It is believed that SARS-COV-2 is able to use angiotensin-converting enzyme 2 (ACE 2) as a receptor for cell entry (Zheng et al., 2020; Zhou et al., 2020b). ACE2 is attached to the outer surface (cell membranes) of cells in the lungs, arteries, heart, kidney, and intestines (Hamming et al., 2004). Additionally, ACE2 is expressed in Leydig cells in the testes (Jiang et al., 2014) and neurological tissue (Baig et al., 2020). As such, it is possible that these organs might also be infected by SARS-CoV-2. The host immune response to SARS-CoV-2 may also impact pathogenicity, resulting in severe tissue damage and, occasionally, death. Indeed, several studies have reported lymphopenia, exhausted lymphocytes and cytokine storms in COVID-19 patients (Blanco-Melo et al., 2020; Cao, 2020). Numerous clinical studies have also observed the elevation of lactate dehydrogenase (LDH), IL-6, troponin I, inflammatory markers and D-dimer in COVID-19 patients (Wang et al., 2020b; Zhou et al., 2020a). However, despite the enormous burden of morbidity and mortality due to COVID-19, we know little about its pathophysiology, even though this establishes the basis for successful clinical practice, vaccine development and drug discovery.

Using a multi-omics approach employing cutting-edge transcriptomic, proteomic and metabolomic technologies we identified significant molecular alterations in patients with COVID-19 compared to uninfected controls in this study. Our results refine the molecular view of COVID-19 pathophysiology associated with disease progression and clinical outcome.

## Results

### Patient cohort and clinical characters

We studied 66 clinically diagnosed and laboratory confirmed COVID-19 patients hospitalized at the Shanghai Public Health Clinical Center, Shanghai, China between January 31st and April 7th, 2020 (Fig. 1A, Tables S1 and S2). At the time of writing, 55 (49 mild and 6 severe) of the 66 patients have recovered and been discharged following treatment, while five patients (1 mild and 4 severe) remain in the hospital and are receiving ongoing treatment. Unfortunately, six patients (all severe) died.

**Fig. 1.**
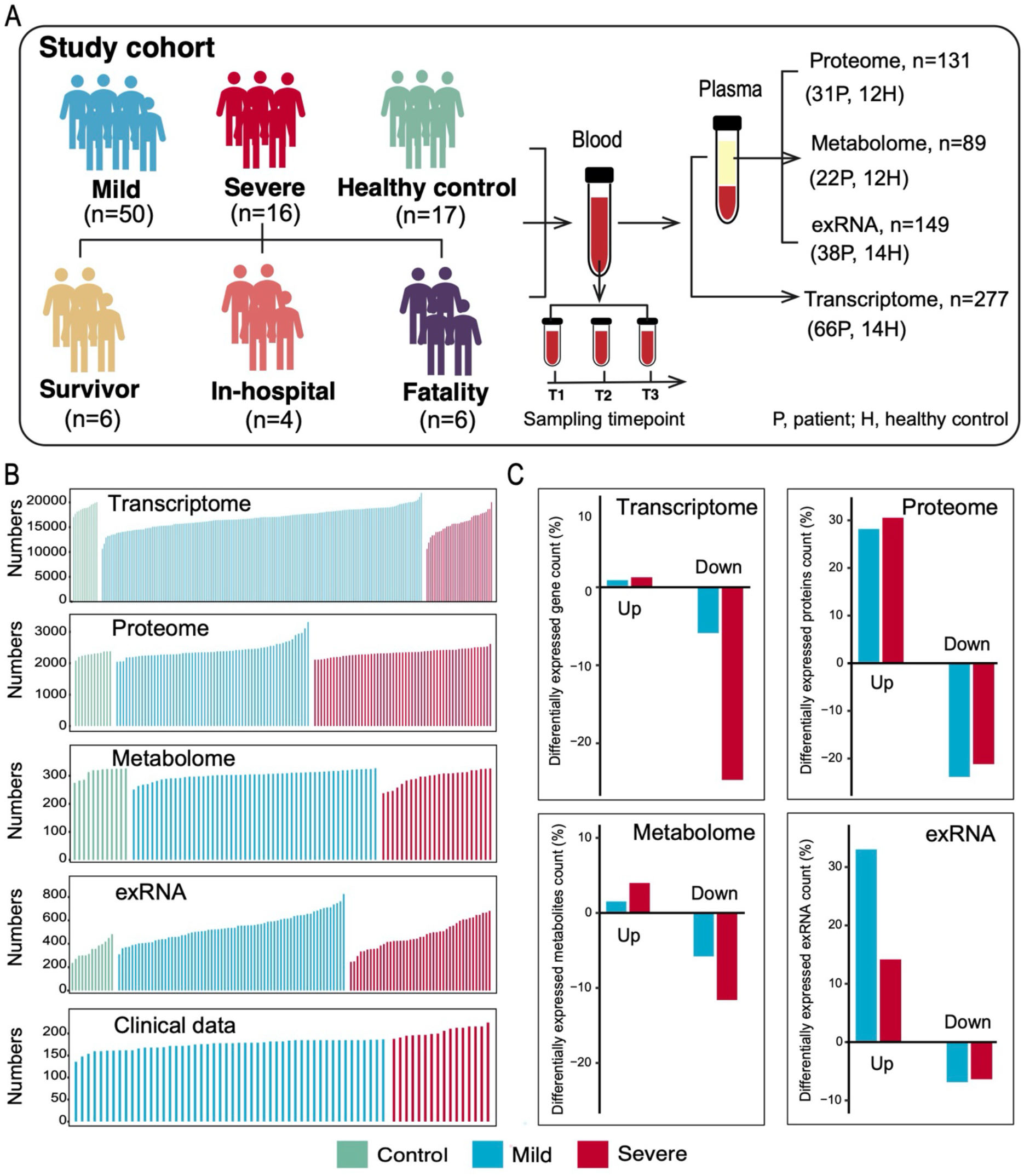
Study design and patient cohort. (A) Schematic summary of the study design and patient cohosrt. Total sample numbers before quality control are shown for each omics data set. (B) The number of expressed genes and detected proteins, metabolites, exRNAs and clinical parameters in high quality patient samples. (C) Summary of differentially expressed genes, proteins, metabolites and exRNAs between uninfected controls and COVID-19 patients (mild and severe) in the multi-omics data.

### Molecular variation associated with COVID-19 pathophysiology

Serial blood and throat swab samples were collected from all patients, as well as from 17 healthy volunteers. To determine whether COVID-19 pathophysiology was associated with particular molecular changes, a total of 23,373 expressed genes, 9,439 proteins, 327 metabolites and 769 exRNAs were examined using a multi-omics approach combining transcriptomics, proteomics, and metabolomics (Fig. 1B). Compared with healthy controls, mild and severe patients had significantly different expression patterns (higher or lower) in 6.79% and 26.0% of expressed genes, 52.1% and 51.7% of proteins, 7.34% and 15.6% of metabolites and 39.9% and 20.5% of exRNAs, respectively (Fig. 1C, Tables S3-S6). Significant differences in the principal component 1 (PC1), PC2 and/or PC3 between healthy controls, mild and severe COVID-19 patients were observed (Figs. 2A and S1A). Remarkably, there were significant correlations between multi-omics data and classical blood and biochemical parameters (Fig. 2B), suggesting that the molecular changes identified directly impact the pathophysiology of COVID-19.

**Fig. 2.**
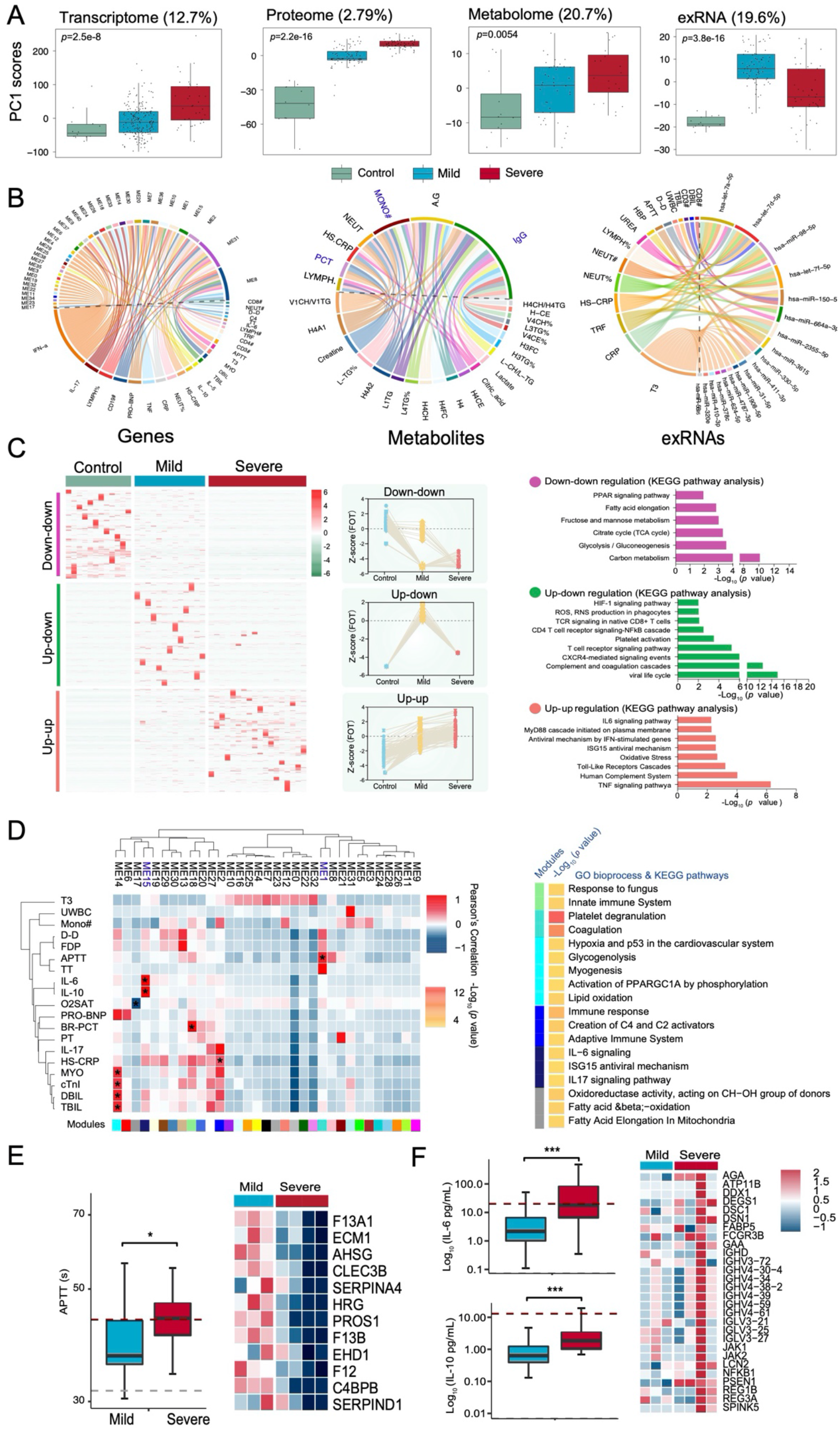
Molecular characteristics of COVID-19 patients. (A) Scores of principal components 1(PC1) of each sample from the transcriptome, proteome, metabolome, and exRNA-seq principal component analyses. (B) Circos plots showing the significant correlations between clinical parameters and the multi-omics data. (C) The cluster heatmap represents expression patterns in the 1,656, 1,547 and 2,362 proteins that showed significant upregulation (fold change>2) in the control, mild and severe patient groups. The top categories enriched for clusters are shown. Values for each protein in all analyzed regions (columns) are color-coded based on expression level, low (blue) and high (red) z-scored FOT. (D) The WGCNA of COVID-19 samples shows modules that are highly correlated with clinical features (left heatmap). Enrichment analysis for different modules is presented in the right heatmap (*p* value <0.05). (E) Boxplot indicates the APTT time between mild and severe COVID-19 patients. The heatmap indicates the module 1 enriched protein expression patterns among mild and severe patients. (F) Boxplot indicates the IL-6 and IL-10 levels among mild and severe COVID-19 patients. The heatmap indicates the module 2 enriched protein expression patterns among mild and severe patients. Differences between groups were estimated using ANOVA. For all boxplots, the horizontal box lines in the boxplots represent the first quartile, the median, and the third quartile. Whiskers denote the range of points within the first quartile − 1.5× the interquartile range and the third quartile + 1.5× the interquartile range.

The correlation between molecular variation and COVID-19 pathophysiology was best reflected in the proteomic analysis (Fig. 2). Specifically, there was significant downregulation in the tricarboxylic acid cycle (TCA) and glycolytic pathways in both mild and severe patients compared to healthy controls (Figs. 2C and S1B). However, the hypoxia-inducible factors (HIF-1) signaling pathways and well-known host defense pathways (e.g., T cell receptor signaling pathway, ISG15 antiviral signaling pathway) were elevated in these patients (Figs. 2C and S1B). Additionally, we identified 14 co-expression groups (“modules”) of proteins that were highly correlated to clinical parameters (Fig. 2D). Module 1, comprising 12 proteins, was strongly associated with activated partial thromboplastin time (APTT) (Fig. 2E), with their downregulated expression likely indicating higher APTT values. In contrast, levels of plasma IL-6 and IL-10 in patients were positively correlated with the expression of proteins in module 15 (Fig. 2F). Notably, correlations between the proteins in these modules were also identified, suggesting that proteins may interact in defining clinical outcome (Figs. S2C and S2D). In addition to proteins, lipoprotein variation was also significantly correlated with immune changes including IgG, monocytes and procalcitonin (Fig. 2B). Combined, these data reveal a significant association between specific molecular variations and COVID-19 pathophysiology.

### Tissue damage caused by SARS-CoV-2

Recent data suggests that SARS-CoV-2 infection is associated with multi-tissue injury and organ damage (Wu et al., 2020b; Zheng et al., 2020).Compared to healthy controls, intensive alteration of tissue-enhanced proteins were observed in all COVID-19 patients, suggestive of multiple organ dysfunction including the lung, liver, brain, testis and intestine (Figs. 3A and 3I). Notably, the majority of proteins related to organ function were downregulated in COVID-19 patients (Fig. 3B). As expected, lung-enhanced proteins varied significantly in the plasma of both mild and severe patients. Likely because lung-enhanced proteins are not rich in the human protein atlas, the lung-enhanced proteins in either mild or severe patients did not achieve the top rank. Nevertheless, lung abnormality was reflected in the activation of the HIF-1 signaling pathway and reactive oxygen species metabolic processes in all patients (Fig. 3D). Liver- and brain-enhanced proteins also varied significantly, followed by those from the testis, intestine and other organs, suggesting that these organs might also be seriously affected (Fig. 3A). Severe brain dysfunction was reflected in the significant decline of brain-enhanced proteins regulating neurotransmitter synthesis, neurotransmitter transport, and the numbers of neurotransmitter receptors, as well as a significant decrease in proteins including ENO1, MBP and NEFM that are known biomarkers to reflect brain dysfunction (Figs. 3C and 3F). Liver-enhanced proteins, that regulate the transportation of sterol and cholesterol, were downregulated, while those involved in acute inflammatory response were elevated in both mild and severe patients (Fig. 3E). Testis-enhanced proteins involved in the cell cycle and cell proliferation were upregulated in all male patients, although proteins (e.g. YBX2) associated with reproduction were significantly downregulated. Heart specific proteins related to cardia muscle contraction and oxidative reduction were reduced in COVID-19 patients (Fig. 3H). Finally, variation in tissue-enhanced proteins was also associated with COVID-19 severity. For example, brain-enhanced proteins enriched in tubulin accumulation were upregulated in mild disease patients, indicating multiple neuron cell apoptosis in these patients. However, proteins significantly upregulated in severe patients were enriched in liver steatosis AOP and in multi-drug resistance factors (Figs. S2A and S2B).

**Fig. 3.**
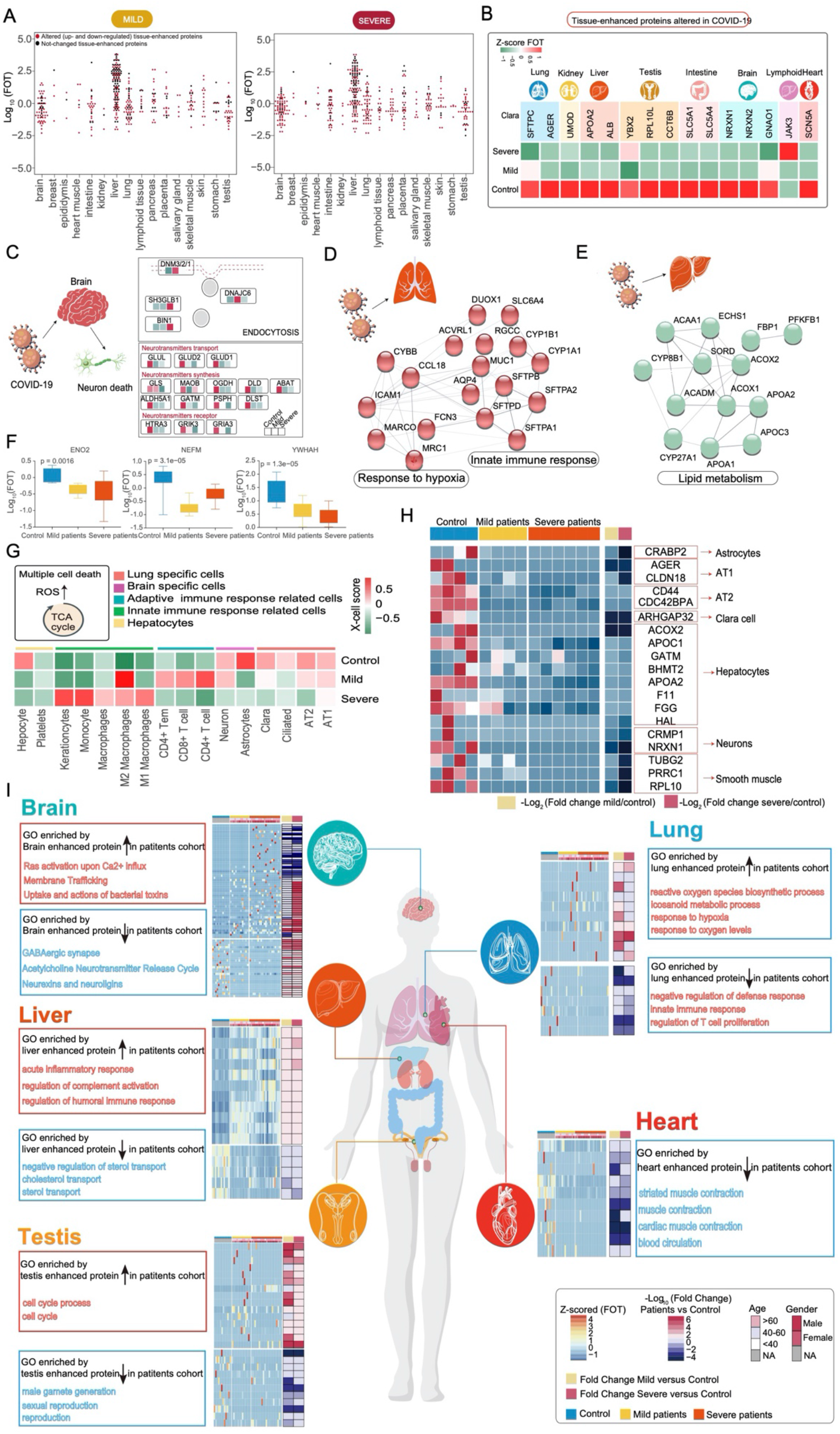
Patterns of tissue damage associated with COVID-19. (A) The distribution of tissue-enhanced proteins across the major tissues affected by COVID-19. Proteins whose expression altered in COVID-19 patients are colored red. (B) Heatmap indicating expression patterns of known tissue specific biomarkers among control, mild and severe patient groups. (C) Systematic summary of brain-enhanced expressed proteins and signaling cascades significantly altered in COVID-19 patients (neurotransmitters transport, synthesis). Values for each protein at all analyzed samples (columns) are color-coded based on the expression level, low (green) and high (red) z-scored FOT. (D) Network summarizing lung-enhanced expressed proteins and signaling cascades significantly altered in COVID-19 patients (HIF-1α signaling pathway). (E) Network summarizing liver-enhanced expressed proteins and signaling cascades significantly altered in COVID-19 patients (Lipid metabolism). (F) Boxplot indicating the expression level of known brain dysfunctional biomarkers in control, mild and severe patients. (G) Heatmap showing normalized x-cell scores of specific cell types across control, mild and severe COVID-19 patients. \**p* < 0.05 (t test). (H) Heatmap showing the expression of cell type specific signatures among control, mild and severe COVID-19 patients. (I) Systematic summary of the GO pathways enriched by tissue-enhanced proteins that exhibited altered expression among control, mild, and severe patient groups. The heatmap of each panel indicates expression patterns of tissues-enhanced proteins among control, mild, and severe patient groups. The fold changes in tissue-enhanced proteins between mild/severe patient samples and control samples are shown on the right of heatmap.

Organ dysfunction was also reflected in the relative proportion of different cell populations. We identified 16 cell types whose abundance changed significantly following virus infection (Fig. 3F). For example, the set of proteins expressed by alveolar type 1/2 epithelial cells (AT1 and AT2) were significantly downregulated in all patients (Fig. 3G). In addition, the majority of tissue-injury related exRNAs across all tissues showed differential expression, including lung (55 in 92 pre-identified, *p*<0.0001), kidney (14 in 22, *p*<0.0001), liver (17 in 22, *p*<0.0001), brain (8 in 16, *p*=0.0016) and heart (5 in 6, *p*<0.0001) (Fig. S2C). Furthermore, a large proportion of tissue-injury related exRNAs were expressed differently in mild and severe patients in most tissues analyzed (30/92 in lung, 10/22 in kidney, 8/22 in liver and 3/6 in heart), except brain (1/16) (data not shown). Together, our data indicate that COVID-19 causes adverse functions in multiple organs including lung damage.

### Immunopathological changes in COVID-19 patients

Immune responses can cause severe damage to the cells or tissues that defend hosts against viral infection (Baseler et al., 2017; Cicchese et al., 2018; Newton et al., 2016). Analysis of whole blood transcriptomic data revealed that gene sets, including an antiviral IFN signature (M75 module), were enriched at the first sampling timepoint (Fig. 4A, Table S7). Notably, IFN signaling was continuously activated in severe patients during the entire period of hospitalization (Fig. 4A), while negative regulators of innate immune signaling (e.g. TRIM59, USP21 and NLRC3) were downregulated (Fig. S3A). Additionally, significant increases of IL-6, IL-8 and IL-10 levels were detected in severe patients compared to mild patients (Figs. 2F and S3B). Combined, these data suggest that the continuous activation of IFN-I signaling and a high level of inflammatory cytokines likely impact COVID-19 immunopathology.

**Fig. 4.**
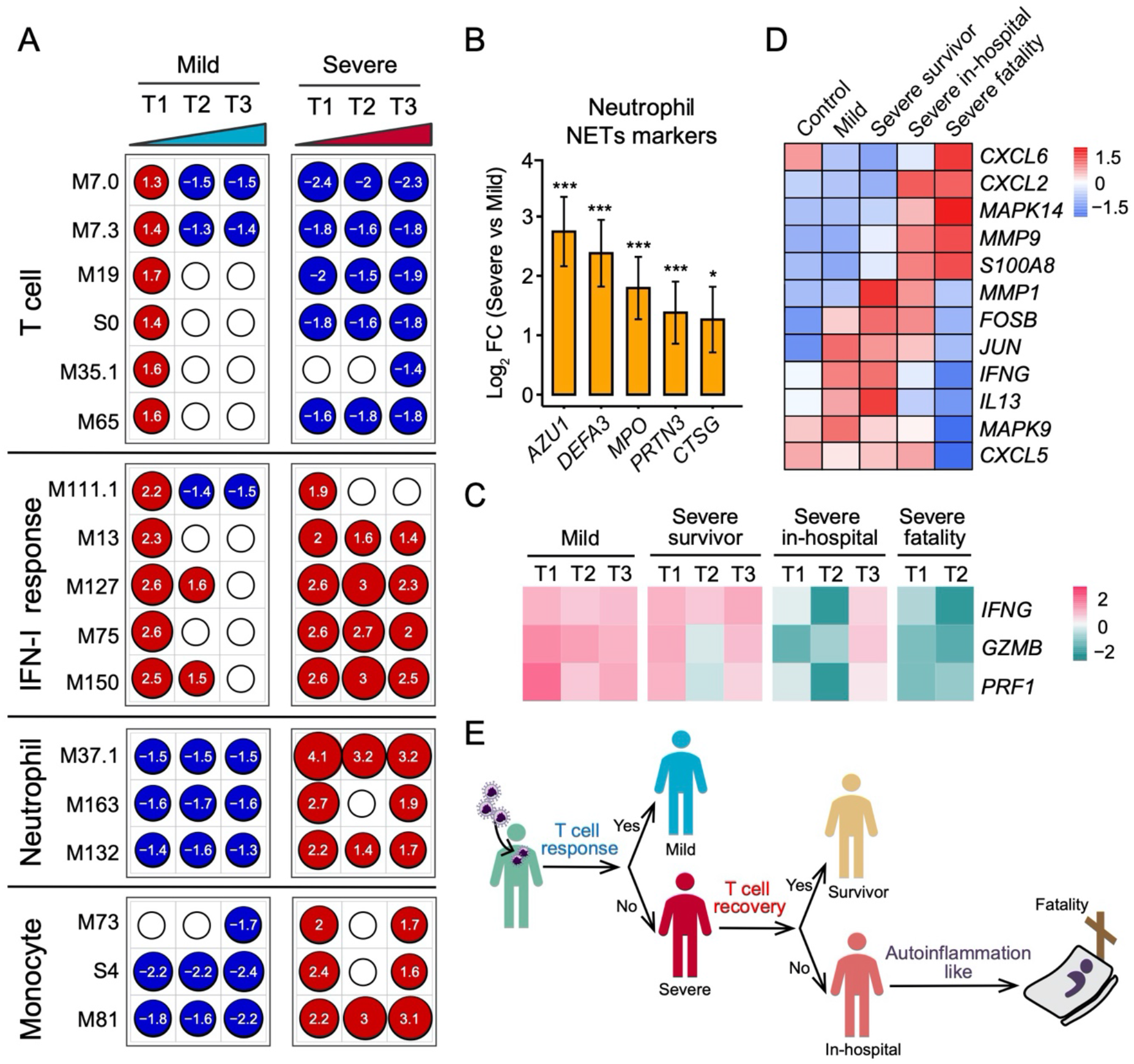
Differences in immune responses among COVID-19 patients. (A) Transcriptional profiles reflect the dynamic immune response in COVID-19. GSEA (FDR < 0.25; 1,000 permutations) was used to identify positive (red), negative (blue) or no (white) enrichment of BTMs (gene sets). The graph shows the normalized enrichment score (NES) of each selected BTM in the different time points (T1, T2 and T3) for patients with mild or severe COVID-19 illness, in comparison to healthy controls. (B) Expression levels of NETs’ markers for individual transcripts in severe versus mild comparisons. Data are represented as means ± SEM, \**p* < 0.05; \*\*\**p* < 0.001 (t test). (C) Heatmap of *IFNG, GZMB* and *PRF1* gene expression in COVID-19 patients. (D) Heatmap of genes enriched in IL-17 signaling pathway between healthy control and COVID-19 patients. (E) T cell and innate immune response elucidate immunopathology of COVID-19.

Higher neutrophil counts were observed in severe patients but not in mild patients during hospitalization (Fig. S3C). Examination of the neutrophil transcriptomic signatures revealed that excessive neutrophil activation was associated with severe rather than mild disease. These markers involved those utilized in neutrophil chemotaxis, activation and migration (Fig. S3D). Notably, genes encoding molecules associated with neutrophil extracellular traps (NETs) were significantly upregulated in severe disease patients (Fig. 4B). As excess NETs formation can lead to tissue damage (Kruger et al., 2015), our data imply that the excessive activation of neutrophils may contribute to COVID-19 pathogenesis.

As in the case of influenza viruses that can be cleared by strong T cell responses (van de Sandt et al., 2014; Wang et al., 2015), SARS-CoV-2 immunity in mild patients was characterized by a robust T cell response, reflected in T cell signaling activation (M7.3 module, M35.1 module) and T cell differentiation (M19 module) on admission, followed by subsequent rapid reduction (Fig. 4A). However, severe patients lost ∼59.1% of their total T cell population, 62.3% of their CD4 T cells and 52.8% of their CD8 T cells. Importantly, the CD4 T cell population gradually recovered in the severe-survivors compared to the severe-fatality group (Fig. S3E). Additionally, T cells in the survivors were primed by dendritic cells and expressed high levels of *IFNG* and *GZMB* (Fig. 4C). T cell dysfunction was observed in the severe group, which could in part be due to an inhibitory status based on the expression levels of multiple exhaustion markers (Fig. S3F). Strikingly, the severe disease group had a greater abundance of *ARG1* (Fig. S3G). Finally, the mild group had higher TCR diversity than the severe group (Fig. S3H). In sum, our data suggest that the T cell response is indispensable to successful host defense against SARS-CoV-2.

Finally, we investigated the immune signatures associated with poor COVID-19 prognosis. Notably, KEGG functional analysis revealed that gene sets of the “IL-17 signaling pathway” were significantly enriched in the severe-fatality group. Further analysis of the signature components revealed that p38 MAPK activation was dominant in fatal cases, while higher levels of IL13 and IFNG were present in survivors (Fig. 4D). These gene signatures might contribute to greater neutrophil influx (CXCL2 and CXCL6) and inflammation (S100A8), and could be detrimental in the severe disease group (Fig. 4E).

### Comprehensive changes in lipoprotein metabolism in COVID-19 patients

To reveal metabolic changes in COVID-19 patients, we quantified 348 metabolite parameters in small metabolites, lipoprotein subclasses and their compositional components. The PCA scores plot revealed an obvious metabolomic trajectory from mild to severe COVID-19, and gradually away from healthy controls (Fig. 5A). Such group-clustering patterns were independently confirmed by PCA scores plots from all NMR-detectable metabolite signals, all MS-detectable signals for lipids, and hydrophilic molecules in plasma samples (Figs. S4A-S4D). Our data therefore indicate that a concentration of changes in plasma metabolites are associated with COVID-19 severity.

**Fig. 5.**
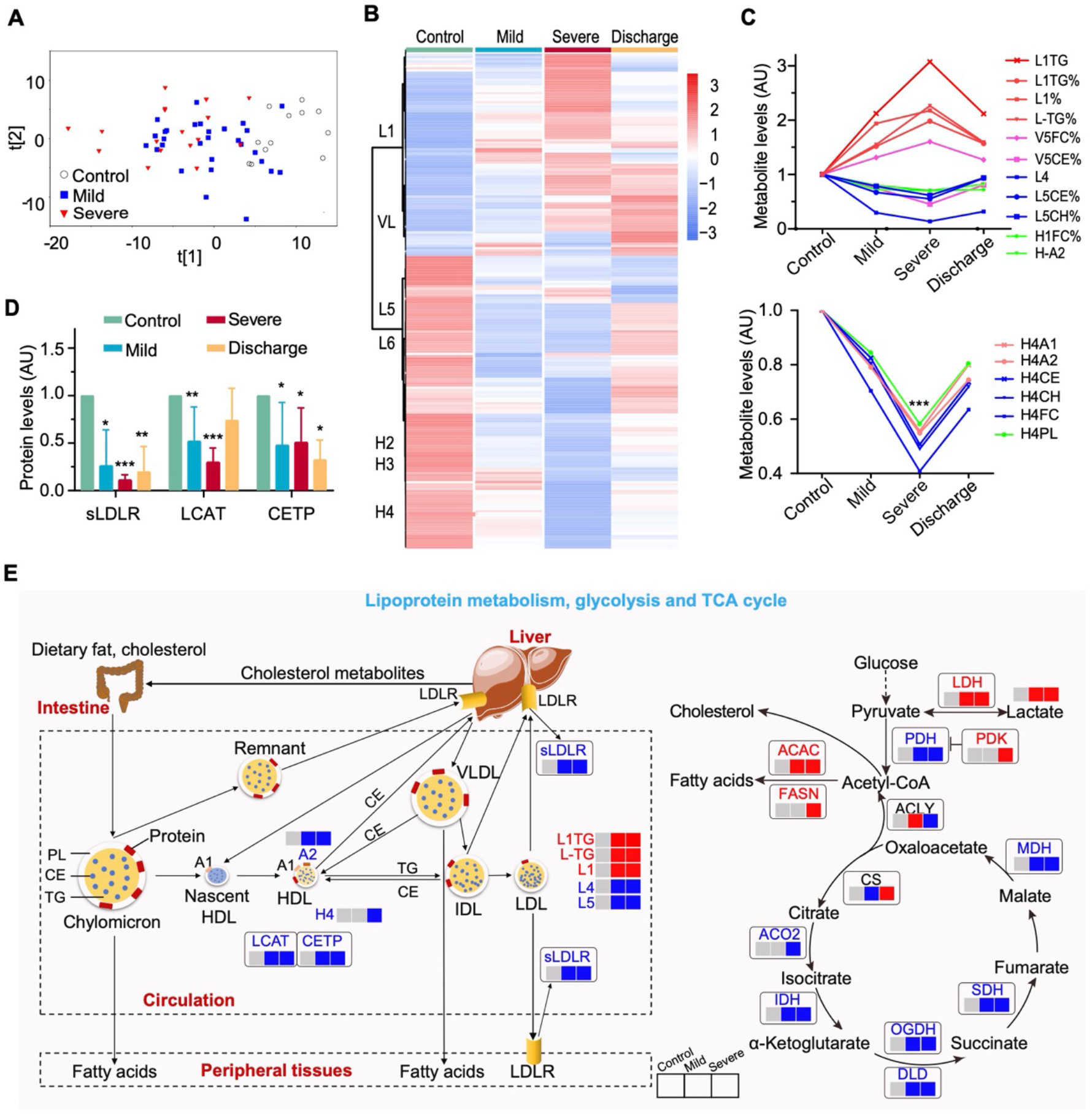
COVID-19-associated metabolomic changes in blood plasma. (A) Plasma metabolomic changes revealed a trajectory in COVID-19 severity, from healthy control, mild, to severe. (B) Changes in the concentration of plasma metabolites are associated with COVID-19 severity. The discharge group consist of all patients (mild and severe) that were recovered and discharged. (C) COVID-19 severity is associated with significant changes in lipoprotein subclasses including high-density lipoprotein subclass-1 (HDL1), HDL4, low-density lipoprotein subclasses (LDL1, LDL4, LDL5), very low-density lipoprotein subclass-5 (VLDL5) and their compositional components (ApoA1, triglycerides, cholesterol). (D) Plasma levels of key enzymes and proteins directly involving lipoprotein metabolism are indicators for COVID-19 severity. \**p* < 0.05; \*\**p* < 0.01; \*\*\**p* < 0.001 (t test). sLDLR: soluble low density lipoprotein receptor; LCAT: lecithin-cholesterol acyltransferase; CEPT: cholesteryl-ester transfer protein. (E) COVID-19 caused dysregulation in lipoprotein metabolism, glycolysis and TCA cycle. The three boxes from left to right are control, mild and severe, in which gray means normal, blue means decrease, red means increase. H1: HDL1; H3: HDL3; H4: HDL4; L1: LDL1, L3: LDL3; L4: LDL4; L5: LDL5; V1: VLDL1; V4: VLDL4; V5: VLDL5; TG: triglycerides; FC: free cholesterol; CE: cholesteryl esters; CH: total cholesterol (i.e., FC + CE); PL: total phospholipids; H4A1, H4A2: ApoA1 and ApoA2 in HDL4; H-A1, H-A2: ApoA1, ApoA2 (in both HDL and nascent HDL); L1TG: TG in LDL1; L1%: LDL1 percentage in all LDL; TG%, FC%, CE%: percentages of TG, FC and CE, respectively, in total lipids of given lipoprotein subclasses; L-TG/L-CH: TG-to-CH ratio in LDL; H4CH/H4TG: CH-to-TG ratio in HDL4; V1CH/V1TG: CH-to-TG ratio in VLDL1. CS: Citrate synthase; IDH: Isocitrate dehydrogenase; ACO2: Aconitase; OGDH: α-ketoglutarate dehydrogenase; DLD: Dihydrolipoyl dehydrogenase; SDH: Succinic dehydrogenase; MDH: Malate dehydrogenase; PDH: Pyruvate dehydrogenase; PDK: Pyruvate dehydrogenase kinase; ACLY: ATP citrate lyase; ACAC: Acetyl coenzyme A carboxylase; FASN: Fatty acid synthetase; LDH: Lactate dehydrogenase.

Further statistical analyses highlighted the major changes in the levels of lipoprotein sub-classes and their compositional components including LDL1 (L1TG), LDL4, VLDL5, HDL1 and HDL4 (Figs. 5C and S4E). Compared with healthy controls, the level of triglycerides (TG) in LDL1 and free cholesterol (FC) in all VLDL5 lipids were significantly elevated in both mild and severe patients, while there were significant decreases in LDL4 and LDL5, cholesterol in LDL, cholesterol esters in VLDL5, Apo-A2 in both HDL and nascent HDL, FC in HDL1 together with total cholesterol and phospholipids (PL). Interestingly, HDL4 and its components had significant lower levels in severe patients. Compared with mild patients, L1TG and PL in HDL1 was increased in severe patients, while cholesterol in HDL1 and HDL2, HDL4 and its components decreased (Fig. S4F). Fortunately, most of these lipoproteins recovered following patients discharge (Figs. 5B, 5C and S4D).

The levels of some key proteins involved in lipoprotein metabolism, including the soluble low-density lipoprotein receptor (sLDLR), lecithin-cholesterol acyltransferase (LCAT) and the cholesteryl-ester transfer protein (CETP), were significantly reduced in mild and severe COVID-19 patients than those in healthy controls (Figs. 5D and 5E). Additionally, enzymes such as ACO2, IDH, OGDH, DLD, SDH and MDH in the TCA cycle were lower in COVID-19 patients compared to healthy controls, while the enzymes central to fatty acid synthesis (Acetyl coenzyme A carboxylase [ACAC] and Fatty acid synthetase [FASN]) were elevated. Finally, significant concurrent elevations in plasma lactate and LDH were observable in patients compared to healthy controls (Figs. 5E, S4G and S4H). In sum, these data reveal the dysregulation in lipoprotein metabolism, glycolysis and TCA cycle during SARS-CoV-2 infection.

### Viral load is associated with disease prognosis of severe COVID-19 patients

The severity and clinical outcome of COVID-19 were also associated with viral load. Overall, SARS-CoV-2 RNA loads on admission were significantly higher in the throat swabs of the five fatal cases compared to those who survived (mean, 1.26×10^5^ vs 3.98×10^3^ copies/mL, respectively; *p* = 0.04) (Fig. 6A). Although viral load declined during the period of hospitalization in both survival and fatal cases, it remained elevated in fatal cases compared to survivors.

**Fig. 6.**
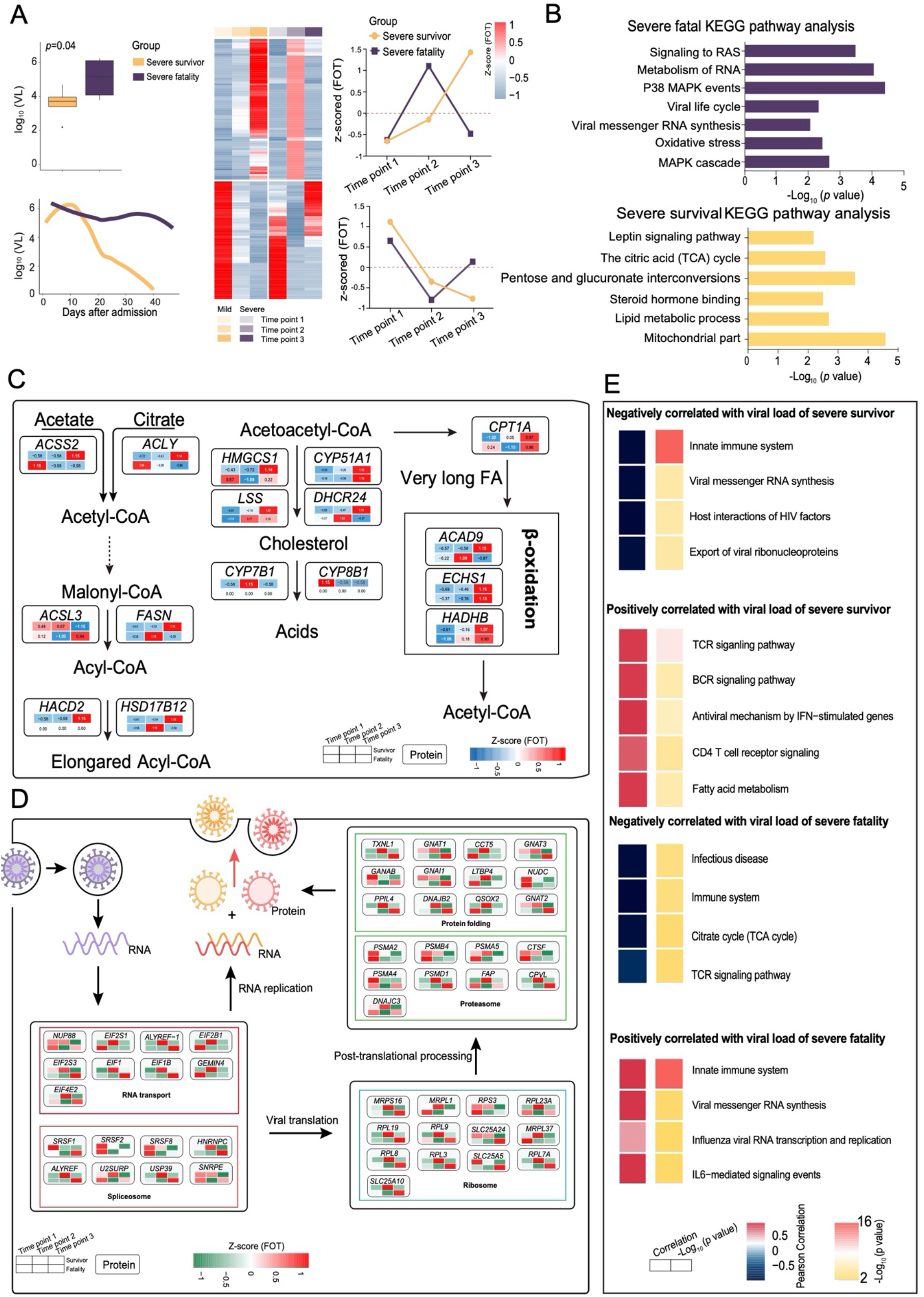
Comparative analysis of severe-survival patients and severe-fatal patients. (A) Bar plot comparison of viral loads between severe-survival patients and severe-fatal patients. The cluster heatmap represents expression patterns of 1,541 proteins that exhibited temporal changes across time points in severe-survival patients. Line graph represents the temporal changes in viral load in time (days) after hospital admission of the patients. (B) Enriched annotations for corresponding clusters showed in Fig. 6A. (C-D) Systematic summary of proteins and signaling cascades significantly altered in severe-survival patients (lipid metabolism; c) and severe-fatal (viral life cycle; d). Values for each protein in all samples analyzed (columns) are color-coded based on the expression level, low (blue) and high (red) z-scored FOT. (E) For each of the four panels, the heatmaps on the left indicated the Pearson correlation of proteins with viral load in severe survivors and severe fatalities, the heatmap on the right indicated the significant pathways enriched by proteins positively or negatively correlated with viral load in severe survivors and severe fatalities.

Estimation of the correlation coefficient between viral load and protein expression revealed that proteins participating in antiviral processes, including the TCR and BCR signaling pathway, were positively associated with viral load changes in severe-survivors. Additionally, proteins participating in viral life cycle processes, including viral messenger RNA synthesis and innate immune responses, were only positively associated with viral load changes in the severe-fatal group (Fig. 6E). Notably, proteins (e.g., FASN, ACSS2, CPT1A, HADHB) involved in pathways including mitochondrial function, lipid metabolic process, steroid hormone process and TCA cycle were continuously upregulated in the severe-survivor compared to the severe-fatal group (Figs. 6B and 6C). However, this upregulation was only observed during the early stage following admission in the severe-fatal group. Surprisingly, proteins related to viral life cycle, viral RNA synthesis, oxidative stress (e.g., EIF, EIFB, RPL19, SLCA24), were downregulated in the several-survivor cases following admission, but maintained high levels in the severe-fatal patients (Figs. 6B and 6D). Hence, SARS-CoV-2 may exploit host resources over the duration of its infection.

### Biomarkers predictive of clinical outcomes of COVID-19 patients

As many molecules associated with COVID-19 pathophysiology were identified, we investigated whether particular molecular changes could be used as biomarkers to predict clinical outcomes. Using an unsupervised PCA, the exRNA, mRNA, proteomics and the corresponding clinical covariate data sets across all time-points, or a subset from the first time-point, clustered into three clinical phenotypes: (i) samples from healthy controls; (ii) samples from COVID-19 patients with a good prognosis; and (iii) samples from COVID-19 patients with a poor prognosis (Methods; Fig. 7A). Given this, prognostic classification models were constructed. Predictive models based on all four types of data worked well, especially those utilizing the clinical covariates and the proteomic data (Fig. S5), suggesting that all four types of data collected at admission contain key prognostic information.

**Fig. 7.**
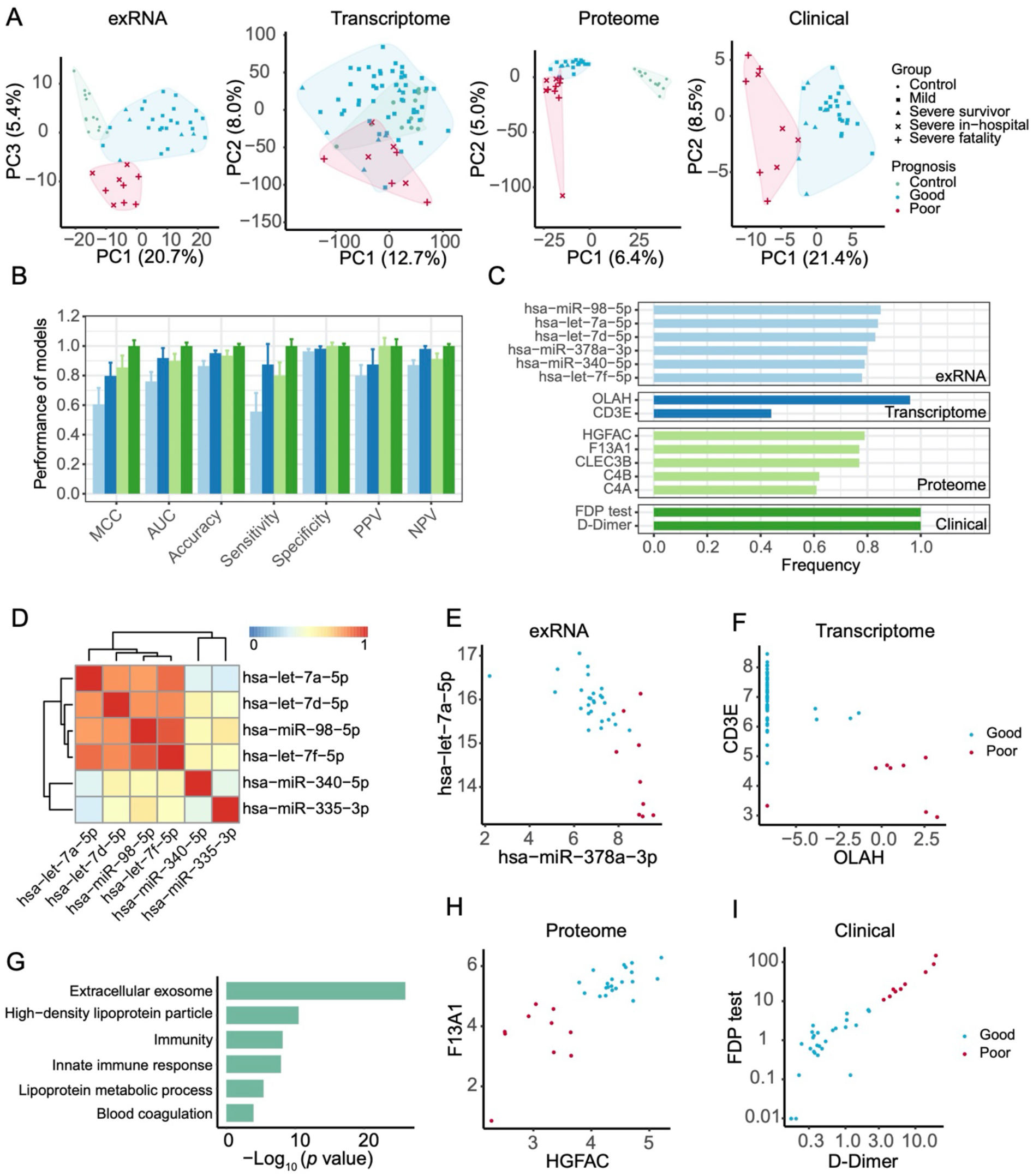
Biomarkers predictive of clinical outcomes of COVID-19 patients. (A) Principal component analysis of exRNA, transcriptome, proteome, and clinical covariate data from samples collected at the first timepoint. The first two components were used to describe the distribution of samples based on expressed genes, proteins and clinical data, respectively, whereas the first and third components were used for samples based on the exRNA data. (B) Performance of prognostic models based on exRNA, transcriptome, proteome, and the corresponding clinical covariate data sets. Model performance of the five-fold cross-validation was assessed using the Matthews correlation coefficient (MCC), AUC, accuracy, sensitivity, specificity, positive predictive value (PPV) and negative predictive value (NPV). (C) The most frequently selected features of exRNA-, transcriptome-, proteome-, and clinical-based models. Features were simultaneously identified from each of the four data sets and for each of the four machine learning algorithms based on the frequency of variables used by AI models during 50 runs of the five-fold cross-validation. (D) Correlation heatmap among the most frequently selected features (frequency > 0.78) used in the exRNA-based model. Members in the let-7 family selected for the exRNA-based predictors (hsa-miR-98-5p, hsa-let-7a-5p, hsa-let-7d-5p, hsa-let-7f-5p) were highly correlated with each other. (E-I) Biomarkers identified from exRNA (E), transcriptome (F), proteome (H) and clinical data (I) based models exhibited a clear separation between those patients with either good or poor prognosis. (G) Functional enrichment of 110 protein features selected from random forest modeling.

In addition, we identified robust predictive models and prognostic biomarkers from each of the four types of data using a previously described approach (Shi et al., 2010) (Figs. 7B and 7C). One or two features (expressed genes, proteins, exRNAs, and biochemical parameters) in each data set were able to clearly separate patients into two groups characterized by different prognoses (Figs. 7D-7I). Poor prognosis was associated with increased levels of D-dimer (*p*=0.004) and fibrinogen degradation products (FDP; *p*=0.02), and with a decrease in F13A1 expression (*p*<0.002; Figs. 7H and 7I), suggesting that blood clotting status may be one of the key factors to monitor in COVID-19 progression. For the mRNA-based model, poor prognosis was associated with lower levels of CD3E and higher levels of OLAH, and hence highly concordant with immune responses in COVID-19 patients (Fig. 7E). Additionally, exRNA-based predictors included members of the let-7 family (Figs.7D and 7E). Finally, the protein-based models highlighted features enriched in extracellular exosomes, lipoprotein metabolic processes, innate immune responses, and blood coagulation (Figs. 7G and 7H).

## Discussion

The COVID-19 pandemic has had a profound impact on a global scale (WHO, 2020). Despite the enormous burden or morbidity and mortality due to COVID-19, we know little about its pathophysiology, even though this establishes the basis for successful clinical practice, vaccine development and drug discovery. Current clinical practice may be unable to provide a precision supportive therapy when a novel disease like COVID-19 emerges, in part explaining the high case fatality rates often observed at the beginning of outbreaks (Alonso et al., 2019). We used a multi-omics approach to identify numerous expressed genes, proteins, metabolites and exRNAs from COVID-19 patients with a range of disease severities, and that were significantly correlated with key clinical features as well as to classic blood and biochemical parameters (Fig. 2). These data therefore provide a comprehensive molecular view of the pathophysiology of COVID-19. Finally, based on our multi-omics data (Fig. S6), mild and severe COVID-19 cases may need different therapeutic strategies.

COVID-19 severity and clinical outcome was significantly associated with multi-organ damage. Due to the widespread presence of ACE2 in humans (Chai et al., 2020; Chen et al., 2020b; Fan et al., 2020; Hamming et al., 2004; Zou et al., 2020), SARS-CoV-2 is able to infect many organs (Wadman et al., 2020). In addition to pneumonia (Wu et al., 2020a; Zhou et al., 2020b), several clinical studies have reported mild gastrointestinal, cardiovascular symptoms and neurological manifestations in hospitalized COVID-19 patients (Baig, 2020; Chen et al., 2020a; Guan et al., 2020; Mao et al., 2020; Xiang et al., 2020; Xu et al., 2020). Histopathologic investigation has also described damage in other organs in addition to the lung (Barton et al., 2020; Cai et al., 2020; Wadman et al., 2020; Wang et al., 2020c). The molecular data generated here support the occurrence of damage in multiple organs including lung, liver, brain, heart in COVID-19 patients, and also identify damage in other organs including the testis (Fig. 3). In the case of brain and testis damage, a key issue is how

SARS-CoV-2 is able to cross the blood-brain or the blood-testis barriers? One possibility might be that heparin was prescribed for coaggregation problems commonly observed in some COVID-19, even though it increases permeability (Gautam et al., 2001; Lin et al., 2020; Oschatz et al., 2011). More importantly, the alteration (up- or down-regulated) of tissue-enhanced proteins and tissue-damage related exRNAs are significantly correlated with clinical severity and outcome.

SARS-CoV-2 infection results in acute lung injury (ALI) in patients, with ground-glass opacity in most computed tomography (CT) reports from our facility and in other hospitals (Chen et al., 2020a; Zhou et al., 2020b; Zhu et al., 2020). Autopsy disclosed histologic changes in lungs included edema, fibrinous/proteinaceous exudates, hyperplastic pneumocytes, patchy inflammation, multinucleated giant cells and diffuse alveolar damage (Barton et al., 2020; Tian et al., 2020). The data generated here revealed that the number of AT1 and AT2 cells reduced significantly in severe patients, suggesting destruction of the alveolar epithelium (Fig. 3F), which in turn will lead to the accumulation alveolar fluid and hence cause hypoxia (Vadász and Sznajder, 2017). In addition, we also noted that HIF1a signaling was modified which may further worsen ALI (Dada et al., 2003). Thus, our molecular data suggest that removal of excess alveolar fluid and the restoration of alveolar structure will be of major clinical importance.

Our data also identified immune pathophysiology a factor that greatly impacted COVID-19 clinical outcome. The innate immune response against viruses is mounted immediately after a host acquires a viral infection, whereas there is a delay before the onset of adaptive immunity (Murphy and Weaver, 2016). Unlike SARS-CoV and influenza virus, SARS-CoV-2 may be present in patients for longer time periods, especially those with severe syndrome (Du et al., 2020; Wang et al., 2020a). Several studies have reported that severe COVID-19 patients experienced lymphopenia, impaired adaptive immunity, uncontrolled inflammatory innate responses, and cytokine storms (Guan et al., 2020; Huang et al., 2020; Qin et al., 2020; Shi et al., 2020; Wang et al., 2020b). While it is believed that T cells play an important role in fighting the infection in the case of Ebola virus, influenza virus and SARS-CoV (Channappanavar et al., 2014; Ruibal et al., 2016; Sridhar et al., 2013; van de Sandt et al., 2014; Zhao et al., 2010), a role for T cells in SARS-CoV-2 infection not yet has been determined, likely reflecting “lymphopenia” (Wang et al., 2020d; Zhou et al., 2020c).

Our longitudinal analyses provided evidence that patients with mild or severe symptoms who succeeded in T cell mobilization promptly controlled SARS-CoV-2 infection and symptoms (Figs. 2C, 4A and 4C). In contrast, those (especially severe-fatal) patients that failed to mount a sound T cell response maintained a continuous pro-inflammatory response and suffered from cytokine storms as well as excess NETs (Figs. 4A and 4B), both of which are known to cause systematic tissue damages (Akiyama et al., 2019; Bohmwald et al., 2019). In sum, our data indicate that T cells play a key role in controlling SARS-CoV-2 infection.

It is believed that p38 signaling, collagenase (MMP9), neutrophil chemo-attractants (CXCL2 and CXCL6) and S100A8 are autoinflammation-like signatures (Cheng et al., 2019; Chung, 2011; Halayko and Ghavami, 2009; Mattos et al., 2002). Remarkably, these molecules were significantly upregulated in severe-fatal in comparison to mild and severe-survival patients (Fig. 4D), the former of which also exhibited a persistent elevation of type I interferon responses (Fig. 4A). Together, the data generated here indicate that autoinflammation may amplify disease in very severe cases of COVID-19.

Patients will receive better and precision therapy if we are able to identify molecular biomarkers associated with prognosis at the beginning of disease presentation. For example, a 21-gene expression assay, which can predict clinical outcome, is used in the case of breast cancer (Sparano et al., 2018). To date, however, almost no biomarkers have been used to accurately predict prognosis in the case of emerging infectious diseases (Wynants et al., 2020). In this study, some of molecules identified at the beginning phase of COVID-19 were significantly correlated to both classical blood and biochemical parameters, and more importantly to disease severity. Based on our previous work (Shi et al., 2010; Su et al., 2014a; Zhang et al., 2015), we established classification models based on each of four data types: exRNAs, mRNA, proteins, and biochemical parameters. Notably, COVID-19 clinical outcomes could be accurately predicted using just one or two biomarkers in each data type. In addition, these biomarkers may have biological functions directly relevant to COVID-19 pathophysiology. For example, biomarkers *let*-7 family from exRNAs, OLAH and CD3E from mRNAs, and C4A and C4B from proteomes concordantly revealed the importance of T-cell activation and the suppression inflammatory responses. However, because of the relatively small patient sample size utilized here, it is clear that more work is needed to confirm the reliability and practicality these biomarkers.

In sum, we have identified a large number of molecules associated with COVID-19 pathophysiology, some of which may also be effective predictive biomarkers of clinical outcome at the onset of disease. In addition, these data suggest that both distinct immune responses and multi-organ damage have a major impact on COVID-19 severity and disease prognosis.

## Materials and Methods

### Study design and patient cohort

According to arrangements made by the Chinese Government, all adult patients in Shanghai diagnosed with COVID-19 were admitted to the Shanghai Public Health Clinical Center. We enrolled 66 COVID-19 patients who were treated at the Shanghai Public Health Clinical Center between January 31^st^ and April 7^th^, 2020. Based on clinical signs and the need for oxygen, these patients were divided into two groups: (i) mild (50/66, 75.8%) – with clinical signs of pneumonia but without oxygen support, and (ii) severe (16/66, 24.2%) – with oxygen support using non-invasive ventilation, tracheal tube, tracheotomy assist ventilation or extracorporeal membrane oxygenation (ECMO) (Fig. 1A and Table S1). All human samples included in the present study were obtained after approval of the research by the Shanghai Public Health Clinical Center Ethics Committee (YJ-2020-S018-02), together with the written informed consent from each patient.

### Sample collection and processing

A total of 277 blood samples, comprising 1-2 mL each, were collected by professional healthcare workers over a 5-week period, with one to five sampling time points from each patient. In addition, 17 blood samples were collected from 17 uninfected volunteers and utilized as healthy controls. Samples were transported to the research laboratory within two hours of collection. For RNA extraction, 200 μL of whole blood was mixed with 1 mL QRIzol reagent (Qiagen), followed by 15 min incubation at room temperature and subsequent freezing at −80°C before total RNA extraction. The remaining whole blood samples (800-1800 μL) were processed immediately to separate plasma and subsequently stored at −80°C until use.

All clinical data were recorded by the clinicians. COVID-19 loads were determined by quantitative real-time RT-PCR using the Takara One Step PrimeScript RT–PCR kit (Takara RR064A) as previously described (Wu et al., 2020a). Quantitative viral load tests were performed using the BioDigital General dPCR kit (Jiangsu Saint Genomics, Cat no. CSJ-3-0018).

### RNA and exRNA extraction and library construction

Total RNA from whole blood samples was extracted using the RNeasy Plus Universal Mini Kit (Qiagen) following the manufacturer’s instructions. The quantity and quality of RNA solution were assessed using a Qubit Flex fluorometer (Invitrogen) and an Agilent Bioanalyzer (Agilent Technologies) before library construction and sequencing. RNA library construction was performed as described using the VAHTS Universal V6 RNA-seq Library Prep Kit for Illumina (Vazyme, China). Ribosomal, globin and RN7S RNAs were depleted using specially designed probes (Vazyme, China).

Plasma samples were divided into aliquots and used for extracellular RNA (exRNA) extraction and library construction, protein extraction and metabolomic analyses. For exRNA library preparation, total RNA including exRNA was extracted using the miRNeasy Serum/Plasma Advanced Kit (Qiagen). The exRNA library was prepared using the NEXTflex Small RNA-seq Kit v3 (PerkinElmer). RNA quantity and quality were determined as mentioned above. After final library quantification using a Qubit Flex fluorometer (Invitrogen) and quality control using the Bioptic Qsep100 to confirm the expected size distributions, all libraries (RNA and exRNA) were pair-end (150-bp reads) sequenced on the Illumina NovaSeq6000 platform (Illumina).

### RNA-seq data analysis

#### Data processing and filtering criteria

Preliminary processing of raw reads was performed using FASTP v0.19.6 to remove adapter sequences and obtain trimmed reads (Chen et al., 2018). The sequence AGATCGGAAGAGCACACGTCTGAACTCCAGTCA was used as the R1 adapter sequence while AGATCGGAAGAGCGTCGTGTAGGGAAAGAGTGT was used as the R2 adapter sequence. HISAT2 v2.1 (Pertea et al., 2016) was used for read alignment to the human genome, build 38. Samtools v1.3.1 was used to generate intermediate result files for quality assessment of the aligned reads by BamQC v2.0.0 (https://github.com/s-andrews/BamQC) and insert size distribution analysis. The assembly of aligned reads and assessment of expression levels were processed through StringTie v1.3.4. Gene counts were determined with preDE.py (http://ccb.jhu.edu/software/stringtie/) based on results derived from Ballgown (https://github.com/alyssafrazee/ballgown). Ensembl transcript annotation (version: Homo_sapiens.GRCh38.93.gtf) with 58,395 genes was used.

A QC analysis and library filtering were performed before downstream biological analysis. Libraries that passed the following criteria were retained: (i) more than five million reads; (ii) more than 90% of reads aligned to the human reference genome; (iii) over 10,000 genes were expressed (a gene with FPKM>0.5 was identified as an expressed gene). In addition, to monitor data quality across batches, libraries of some heathy control samples were constructed and sequenced 2-3 times. The average expression profile of the multiple libraries from each heathy control sample were calculated for follow-up analyses.

#### Immunoassay

Immune repertoires were extracted with MiXCR, a software tool that extracts T-cell receptor (TCR) and immunoglobulin (IG) repertoires from RNA-seq data (Bolotin et al., 2017; Bolotin et al., 2015). The number of clonotypes was then calculated using VDJtools, using the output from MiXCR (Shugay et al., 2015).

#### Differentially expressed genes (DEGs)

To identify DEGs, a Student’s t-test was applied to the expression matrix. Genes with p-values less than 0.05 as well as a fold change >2 or <1/2 were labeled as up-regulated and down-regulated genes, respectively (Su et al., 2014b). This straightforward approach of combining a fold change cut-off with a non-stringent p-value threshold has been demonstrated to yield reproducible and robust lists of DEGs for both microarray and RNA-seq based gene-expression analyses (Shi et al., 2006; Su et al., 2014b).

#### Functional and cell type enrichment analyses

Functional analyses were conducted based on genes differentially expressed between several subgroups of COVID-19 patients compared with healthy control samples. GSEA (Gene Set Enrichment Analyses) was performed to identify significantly enriched functional classes of gene sets correlated with blood transcription modules (BTM) described by Li et al. (2014), KEGG pathways, and Gene Ontology (GO) terms. A default FDR (false discovery rate) value of q□<□0.25 was considered statistically significant. The Normalized Enrichment Score (NES) of significant immune modules from BTMs was used to denote enrichment levels.

The fraction of the cell subsets was calculated using the enrichment-score based algorithm xCell from the RNA-seq data (Aran et al., 2017). Briefly, the expression profile (FPKM) of all 230 samples was employed as raw signatures. The R package immunedeconv was applied to obtain enrichment scores of 35 immune cell types, estimating immune cell fractions including T cell, monocyte and neutrophil by summation of the scores in each sample (Sturm et al., 2019).

### exRNA-seq data analysis

#### Alignment, quantification and quality control

Libraries were sequenced in two batches, with an average sequencing depth of 15.7M raw reads per library. All FASTQ files were delivered to the ExceRpt small RNA sequencing data analysis pipeline (docker v4.6.3) (Rozowsky et al., 2019). Default parameters were used with exception of: (i) the sequence TGGAATTCTCGGGTGCCAAGG was given as the 3’adapter sequence, ignoring the adapter sequences guessed by the pipeline; (ii) the random barcode length was set to 4; (iii) the priority of the reference libraries during read assignment was set to miRNA > piRNA > tRNA > GENCODE > circRNA (Godoy et al., 2018). Pre-compiled genome and transcriptome indices of human genome, build 38 were used. The raw read count matrix was then normalized using count per million (CPM).

A QC analysis was performed prior to biological analysis by removing (i) libraries with low sequencing depths (<1M raw reads); (ii) libraries with mapping ratio lower than 50%, and (iii) libraries with low transcript-genome ratios. To minimize the impact of noise due to low expression levels, only 769 miRNAs with at least 1 count per million in no less than 10% of the total number of samples were included in the final analysis.

#### Differentially expressed exRNAs

To identify differentially expressed exRNAs, Student’s t-tests were applied to the normalized expression matrix. exRNAs with p-values less than 0.05, as well as fold change > 2 or < 1/2, were labeled as up-regulated and down-regulated exRNAs, respectively, in a similar manner to the RNA-seq data.

#### Tissue-damage related miRNAs

exRNAs reported to be associated with tissue damage were collected from publications (Atif and Hicks, 2019; Godwin et al., 2010; Wang et al., 2010; Wang et al., 2009; Zhou et al., 2016)(Table S8). Fisher’s exact test was used to determine whether the proportions of differentially expressed (DE) exRNAs in tissue-damage related exRNAs were significantly higher than the proportions of DE exRNAs in the entire data set of 769 miRNAs.

### Proteome analysis

#### Plasma Protein extraction and trypsin digestion

Plasma samples used for protein extraction were first removed the tip 14 highest abundance plasma proteins using an immunodepleting kit (Thermo Fisher) according to the manufacturer’s instructions, and then inactivated at 85°C for 10 mins. The depleted plasma was digested by trypsin at an enzyme to protein mass ratio of 1:25 overnight at 37°C, and the peptides were then extracted and dried (SpeedVac, Eppendorf).

#### LC-MS/MS Acquisition of Plasma Samples

Samples were measured using LC-MS instrumentation consisting of an EASY-nLC 1200 ultra-high-pressure system (Thermo Fisher Scientific) coupled via a nano-electrospray ion source (Thermo Fisher Scientific) to a Fusion Lumos Orbitrap (Thermo Fisher Scientific). The peptides were dissolved with 12 μl loading buffer (0.1% formic acid in water), and 5 μl was loaded onto a 100 μm I.D. × 2.5 cm, C18 trap column at a maximum pressure 280 bar with 14 μl solvent A (0.1% formic acid in water). Peptides were separated on 150 μm I.D. × 15 cm column (C18, 1.9μm, 120 A□, Dr. Maisch GmbH) with a linear 15–30% Mobile Phase B (ACN and 0.1% formic acid) at 600 nl/min for 75 mins. The MS analysis was performed in a data-independent manner. The DIA method consisted of MS1 scan from 300-1400 m/z at 60k resolution (AGC target 4e5 or 50 ms). Then 30 DIA segments were acquired at 15k resolution with an AGC target 5e4 or 22 ms for maximal injection time. The setting “inject ions for all available parallelizable time” was enabled. HCD fragmentation was set to normalized collision energy of 30%. The spectra were recorded in profile mode. The default charge state for the MS2 was set to 3.

#### Peptide identification and protein quantification

All data were processed using Firmiana (Feng et al., 2017).The DDA data were search against UniProt human protein database (updated on 2019.12.17, 20406 entries) using FragPipe (v12.1) with MSFragger (2.2) (Kong et al., 2017). The mass tolerances were 20 ppm for precursor and 50 mmu for product ions. Up to two missed cleavages were allowed. The search engine set cysteine carbamidomethylation as a fixed modification and N-acetylation and oxidation of methionine as variable modifications. Precursor ion score charges were limited to +2, +3, and +4. The data were also searched against a decoy database so that protein identifications were accepted at a false discovery rate (FDR) of 1%. The results of DDA data were combined into spectra libraries using SpectraST software. A total of 327 libraries were used as reference spectra libraries.

DIA data was analyzed using DIANN (v1.7.0) (Demichev et al., 2020). The default settings were used for DIA-NN (Precursor FDR: 5%, Log lev: 1, Mass accuracy: 20 ppm, MS1 accuracy: 10 ppm, Scan window: 30, Implicit protein group: genes, Quantification strategy: robust LC (high accuracy). Quantification of identified peptides was calculated as the average of chromatographic fragment ion peak areas across all reference spectra libraries. Label-free protein quantifications were calculated using a label-free, intensity-based absolute quantification (iBAQ) approach (Zhang et al., 2012). We calculated the peak area values as parts of corresponding proteins. The fraction of total (FOT) was used to represent the normalized abundance of a particular protein across samples. FOT was defined as a protein’s iBAQ divided by the total iBAQ of all identified proteins within a sample. The FOT values were multiplied by 10^5^ for the ease of presentation and missing values were imputed with 10^−5^.

### Metabolome analysis

#### NMR spectroscopy

The plasma samples used for NMR analysis were first treated with 56°C for 30 min. Our subsequent quantitative measurements of samples from healthy controls showed that such treatments caused no differences to the quantification results.

NMR analysis was conducted on a 600 MHz NMR spectrometer (Bruker Biospin) as reported previously (Jimenez et al., 2018) with some minor modifications. In brief, 320μL of each plasma sample was mixed with 320μL of a phosphate buffer (0.085 M containing 10% D_2_O) with composition described previously (Jiang et al., 2012), and 600μL mixture was transferred into a 5mm NMR tube for NMR analysis. 152 parameters of the plasma were then quantified using a server-based software package (Bruker Biospin), including 112 parameters for lipoproteins (including main fractions, subclasses and compositional components therein), two acute-phase glycoproteins together with 41 small metabolites (such as amino acids, ketone bodies, glucose, carboxylic acids, ethanol). We also quantified six ratio-parameters for saturated, unsaturated, monounsaturated and polyunsaturated fatty acids from the diffusion-edited spectra (Xu et al., 2012). We further calculated 187 more ratio-parameters (such as the cholesterol-to-triglyceride ratio, percentage of triglycerides and cholesterol in total lipids) from the quantitative data for lipoproteins. A total of 348 quantitative parameters obtained were collectively employed to define the metabolomic phenotypes of each of the human plasma samples.

### Development of prognostic models

#### Model development

Four data sets representing the (i) clinical tests, (ii) exRNA-seq, (iii) mRNA-seq, and (iv) proteomics quantification analysis were used to develop prognostic models for the prediction of patient outcomes (i.e. good or poor). Patients with a “good” outcome included those with mild or severe syndrome but who were discharged after treatment; while patients with “poor” outcomes included those who died or remained in ICU for more than two months.

Prognostic models were developed and validated using a two-layer validation strategy (Fig. S5A) to prevent information leaking from the training set to the validation set^13^. Briefly, patients were first divided into training and validation sets with equal size based on outcome and admission date. The training set was then used to select variables and train prognostic models using multiple machine learning algorithms, including nearest mean classification (NMC), k-nearest neighbors (KNN), support vector machine (SVM), and random forest (RF) through an internal-layer of 50 runs of five-fold cross-validation process to resist overfitting. Next, a final model was built using the whole training set with the best performing machine learning algorithm as defined above. The final model was further validated using the validation set as an external-layer evaluation. Model performance was assessed in terms of the Matthews correlation coefficient (MCC), AUC, accuracy, sensitivity, specificity, positive predictive value (PPV) and negative predictive value (NPV).

Prognostic biomarkers were identified based on the frequency of variables selected by machine learning algorithms. Because the sample size was relatively small compared to the large number of variables, it was difficult to identify stable biomarkers as indicated by the low frequencies of the variables used in the prognostic models^13^ (Figs. S5A and S5B). To detect more robust prognostic biomarkers, 50 runs of five-fold cross-validation process were therefore applied to the whole data set. The variables used by the best performing machine learning algorithm were identified as prognostic biomarkers for each data set.

#### Learning curve model comparison

Learning curve model comparison (LCMC) was performed using Predictive Modeling Review as available in JMP Genomics 10 (https://www.jmp.com/en_us/software/genomics-data-analysis-software.html). LCMC reveals the effects of sample size on the accuracy and variability of the predictive models using 10 runs of 4-fold cross-validation.

We performed LCMC with prognosis (good or poor) as target variables, and the clinical variables, exRNA, mRNA, or proteomics measurements as predictors. Fig. S5C shows each individual (RMSE) and (AUC) learning curve and the average for each of the eight partition tree models for clinical endpoints, as well as exRNA using K-fold cross validation. The LCMC suggested that with up to 15 samples, eight partition tree models reached AUC as 1 for clinical variables. However, more than 23 and 30 samples were needed for one and three models, respectively, to reach AUC of 1 for exRNA-seq data. The variability of RMSE and AUC for the proteomic and mRNA-seq data (not shown) were between that observed for clinical variables and the exRNA data.

### Statistical Analyses

Univariate statistical analysis was performed using student’s t-test, Mann-Whitney U tests or ANOVA tests to compare continuous variables. Chi-square tests and Fisher’s exact tests were used for the comparison of categorical variables. *p*-values were adjusted using Bonferroni correction or the Benjamini and Hochbery False Discovery Rate (FDR) in multiple comparisons, with *p* < 0.05 considered to be statistically significant. Principal components analysis (PCA) was conducted with univariance scaling, with the scores plot showing a distribution of metabolomic phenotypes for healthy participants and patients with moderate or severe COVID-19 (and upon discharge). Correlations were tested using Pearson correlation coefficients. Locally Weighted Linear Regression (Loess) was used for visualizing the time series data. All analyses were performed using appropriate R packages (version 3.5.1).

## Data Availability

The datasets produced in this study are available in the following databases: RNA-Seq and exRNA-Seq Data: NODE OEP000868; Raw mass spectrometry data: iProX IPX 0002186001

## Acknowledgements

We acknowledge the support from Volvo Car Corporation. This study was supported by the National Natural Science Foundation of China (grants 81861138003, 31930001, 32041004 and 81672057) and the Special National Project on investigation of basic resources of China (grant 2019FY101400), and Shanghai Municipal Science and Technology Major Project (Grant No. 2017SHZDZX01). E.C.H. is supported by an ARC Australian Laureate Fellowship (FL170100022).

## Author contributions

Y.-Z.Z. conceived and designed the study. Y.-Z.Z., T.-Y.Z., C.D., L.M.S., H.R.T., Y.L., Z.Y. and L.J. designed and developed methodology. T.-Y.Z., Y.L. and Z.-G.S. performed the clinical work and sample collection. Y.-M.C, Y.-T.Z., Y.-Z.W., Q.-X.H., S.-Y.S., Z.-G.S., F.-H.D., Y.-P.A., Q.-S.C. and Q.-W. L. performed the experiments. Y.Y., Y.-Z.W., Q.-X.H., Y.-M.C., F.Q, L.S., Z.-Y.C., J.-C.Y., Z.-Q.S., J.-W.F. and P.-C.L. analysed the data. Y.-Z.Z., C.D., F.Q, L.S., Y.Y., Y.-Z.W., Y.-M.C., Q.-X.H., L.M.S., H.R.T. and E.C.H. wrote the paper with input from all authors.

## Data Availability

The datasets produced in this study are available in the following databases:

- RNA-Seq and exRNA-Seq Data: NODE **OEP000868** (http://www.biosino.org/node/project/detail/OEP000868)
- Raw mass spectrometry data: iProX **IPX 0002186001** (https://www.iprox.org/page/subproject.html?id=IPX0002186001)

